# SARS-CoV-2 receptor networks in diabetic and COVID-19 associated kidney disease

**DOI:** 10.1101/2020.05.09.20096511

**Authors:** Rajasree Menon, Edgar A. Otto, Rachel Sealfon, Viji Nair, Aaron K. Wong, Chandra L. Theesfeld, Xi Chen, Yuan Wang, Avinash S. Boppana, Jinghui Luo, Yingbao Yang, Peter M. Kasson, Jennifer A. Schaub, Celine C. Berthier, Sean Eddy, Chrysta C. Lienczewski, Bradley Godfrey, Susan L. Dagenais, Ryann Sohaney, John Hartman, Damian Fermin, Lalita Subramanian, Helen C. Looker, Jennifer L. Harder, Laura H. Mariani, Jeffrey B. Hodgin, Jonathan Z. Sexton, Christiane E. Wobus, Abhijit S. Naik, Robert G. Nelson, Olga G. Troyanskaya, Matthias Kretzler

## Abstract

COVID-19 morbidity and mortality is increased in patients with diabetes and kidney disease via unknown mechanisms. SARS-CoV-2 uses angiotensin-converting enzyme 2 (ACE2) for entry into host cells. Since ACE2 is a susceptibility factor for infection, we investigated how diabetic kidney disease (DKD) and medications alter ACE2 receptor expression in kidneys. Single cell RNA profiling of healthy living donor (LD) and DKD kidney biopsies revealed ACE2 expression primarily in proximal tubular epithelial cells (PTEC). This cell specific localization was confirmed by *in situ* hybridization. ACE2 expression levels were unaltered by exposures to renin angiotensin aldosterone system inhibitors in DKD. Bayesian integrative analysis of a large compendium of public -omics datasets identified molecular network modules induced in ACE2-expressing PTEC in DKD (searchable at hb.flatironinstitute.org/covid-kidney) that were linked to viral entry, immune activation, endomembrane reorganization, and RNA processing. The DKD ACE2-positive PTEC module overlapped with expression patterns seen in SARS-CoV-2 infected cells. Similar cellular programs were seen in ACE2-positive PTEC obtained from urine samples of 13 COVID-19 patients who were hospitalized, suggesting a consistent ACE2-coregulated PTEC expression program that may interact with the SARS-CoV-2 infection processes. Thus SARS-CoV-2 receptor networks can seed further research into risk stratification and therapeutic strategies for COVID-19 related kidney damage.

**Translational statement:** To understand the overwhelming burden of kidney disease in COVID-19, we mapped the expression of the SARS-CoV-2 receptor, ACE2, in healthy kidney, early diabetic (DKD) and COVID-19 associated kidney diseases. Single cell RNA sequencing of 111035 cells identified ACE2 predominantly in proximal tubular epithelial cells. ACE2 upregulation was observed in DKD, but was not associated with RAAS inhibition, arguing *against* an increased risk of COVID-19 among patients taking RAAS inhibitors. Molecular network analysis linked ACE2 expression to innate immune response and viral entry machinery, thereby revealing potential therapeutic strategies against COVID-19.

## Introduction

COVID-19 disproportionally affects individuals with diabetes, hypertension, and kidney disease^1-5^. Yet the underlying molecular and physiological causes of this association are unknown, and could be as varied as drugs used to treat these conditions, disease biology^6-8^, direct infection of relevant organs by the virus^9-11^ and consequent tissue destruction, and cytokine storm that occurs secondary to infection^3^. Upper and lower airway tissues are likely the primary sites of infection, however, viral tropism is not restricted to the airways and recent data indicate this includes kidney tissue^10, 12^. COVID-19 patients with kidney disease suffer significantly higher mortality than age-matched individuals without these conditions ^10, 13-15^. Understanding the disease-specific molecular processes associated with COVID-19 in patients with kidney disease and diabetes can have a significant impact on public health.

COVID-19 develops from infection with SARS-CoV-2, a betacoronavirus with a single-stranded RNA genome. It gains entry into specific cell types through interaction of the viral surface spike protein with a cell surface receptor^16^. Studies of severe acute respiratory syndrome (SARS) in the early 2000s identified angiotensin-converting enzyme 2 (ACE2) as the primary cell-entry receptor for the SARS coronavirus (SARS-CoV) in humans^17, 18^. Recent studies demonstrate that ACE2 is also the primary cell-entry receptor for SARS-CoV-2^16, 19, 20^ and, similar to SARS-CoV, ACE2 expression levels correlate with higher risk of SARS-CoV-2 infection^9, 21, 22^.

ACE2 is a membrane-bound zinc metallopeptidase and a master regulator of the renin-angiotensin aldosterone system (RAAS). Normally, ACE2 is expressed as a cell-surface protein to which both SARS-CoV and SARS-CoV-2 can bind^23, 24^. Cleavage of spike protein is required for fusion of the virus with host membranes. The protease TMPRSS2 appears to be the primary protease responsible for this cleavage, at least in lung epithelial cells^25^. However, SARS-CoV-2 can also be internalized by endocytosis and cleavage of the S protein in the endosomal compartment by acid-activated proteases such as cathepsin L (CTSL) and dipeptidyl peptidase-4 (DPP4), both involved in glucose metabolism and immune systems ^8, 20, 26-28^.

Single cell RNA sequencing (scRNAseq) of SARS-CoV-2 target tissues provides a way to identify specific cell types with enhanced ACE2 expression and determine whether these cells possess molecular machinery that facilitate the initial viral entry and subsequent virus-induced cytotoxicity. Characterization of these molecular processes in existing tissue from relevant cohorts could greatly accelerate the identification and development of novel therapeutic options for SARS-CoV-2 infection and non-invasive means for stratifying individuals at risk for COVID-19. Therefore, in this study, we explored the expression and associated biological processes of ACE2 and other SARS-CoV-2 entry factors in kidney cells from healthy living donors (LD), patients with diabetic kidney disease (DKD), and COVID-19 patients that required hospitalization (COV). Using *in situ* hybridization and scRNAseq techniques, we i.) localized the predominant cellular expression of ACE2 in kidneys, ii.) characterized the cellular programs associated with ACE2 regulation in DKD and COV, iii.) mapped the ACE2-associated changes to emerging data on SARS-CoV-2 induced cellular responses, and iv.) tested whether there are associations between disease severity and RAAS medication exposure. Our study included a DKD cohort since COVID-19 disproportionally affects individuals with diabetes and kidney disease^3, 5 8 14, 29, 30^ Side by side analysis of the DKD and COV cohorts aimed to identify ACE2 associated mechanisms in DKD, able to influence pathways also activated in COVID-19 associated kidney disease.

## Results

We studied ACE2 expression in kidney tissue using *in situ* hybridization (ISH) and scRNAseq. We then investigate viral infection associated processes through scRNAseq and functional network analysis (Figure 1).

**Figure 1.**
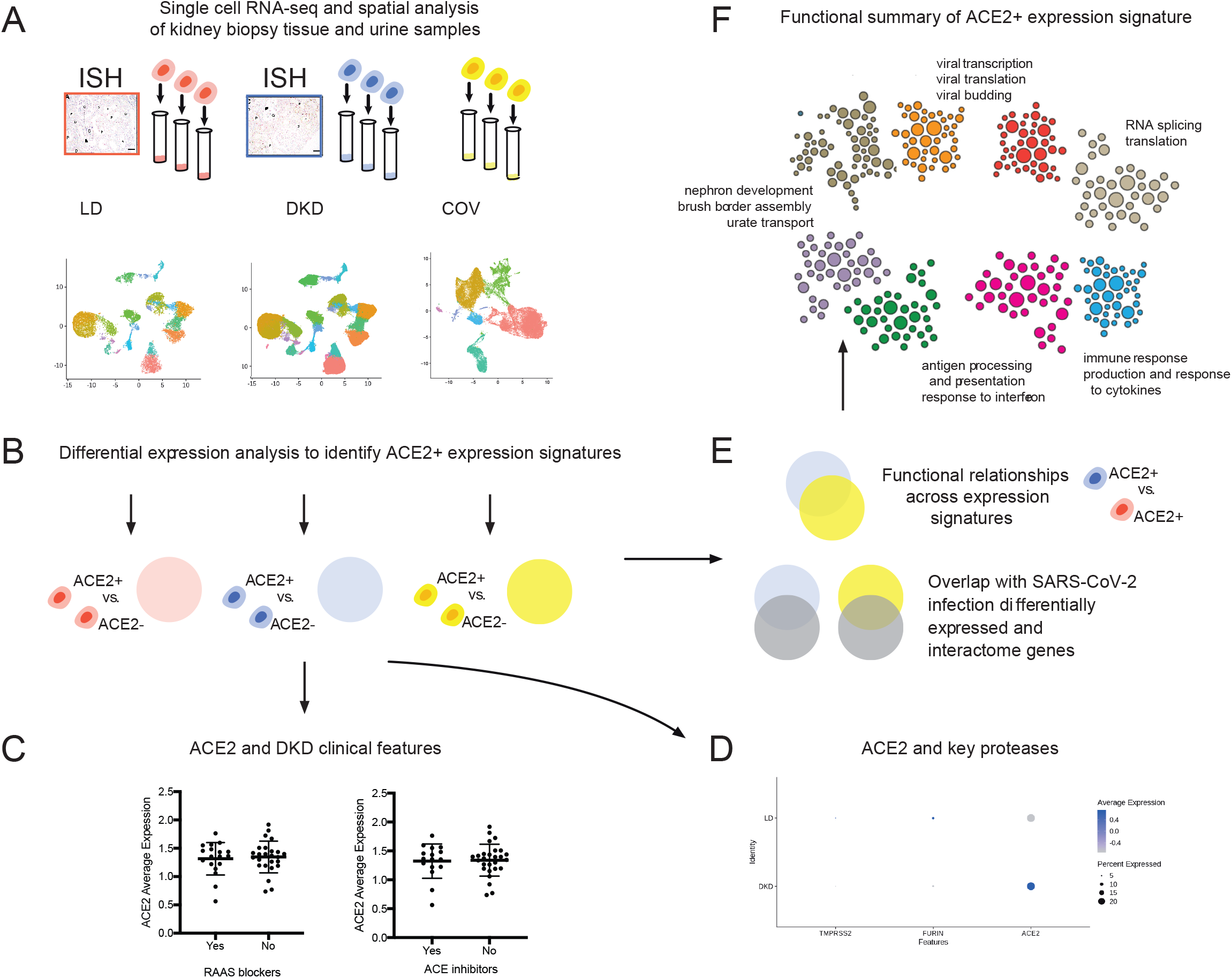
Study overview. To understand how the kidney may be impacted by COVID-19 we performed spatial, systems, and clinical association analyses of ACE2 and other SARS-CoV-2 host factors in kidney biopsies from living donors (LD) and diabetic kidney disease (DKD) patients and kidney cells isolated from the urine of hospitalized COVID-19 patients (COV). (A) Biopsy samples from DKD and living donors (LD) were processed for *in situ* hybridization and scRNAseq profiling. scRNAseq of LD, DKD and urine cell pellets from COV samples were analyzed to determine cell type expression specificity of ACE2 in healthy and disease states. (B) For each scRNAseq dataset, ACE2+ differential expression signatures were identified, (C) Association of ACE2 expression levels in DKD with clinical characteristics were evaluated, including exposure to RAAS blockers and ACE inhibitors. (D) Expression of ACE2 and key proteases between LD and DKD proximal tubule epithelial cells (PTEC) were compared. (E) ACE2 expression signatures across datasets identified aspects induced in PTEC expressing DKD samples compared to LD and found these gene sets to significantly overlap those reported to be impacted by direct SARS-CoV-2 infection. (F) The biological processes in ACE2+ expression signatures were characterized by projecting these signature genes onto PTEC-specific functional networks at HumanBase (hb.flatironinstitute.org/covid-kidney). These networks represent genes and their interactions in biological processes and pathways active in PTEC.

### Cohort studied for SARS-CoV-2 receptor expression and regulation

Kidney cell expression profiles were obtained from early DKD (n=44) and LD (n=18) kidney biopsies and urine samples from COV patients (n=13). Clinical characteristics for the DKD cohort at the time of sample collection are provided in Table 1 and for COV in Supplementary Table 1, with further details in methods.

**Table 1.** Clinical characteristics of diabetic kidney disease cohort at the time of biopsy.

| Characteristic | At time of biopsy<br>(N=44) |
| --- | --- |
| Age, years | 41 ± 11 |
| Diabetes duration, years | 12.2 ± 7.5 |
| Male Sex, n (%) | 14 (32) |
| Body Mass Index (kg/m<sup>2</sup>) | 36.9 (7.3) |
| HbA1c (%) | 9.2 ± 2.4 |
| Systolic blood pressure (mmHg) | 119 ± 12 |
| Diastolic blood pressure (mmHg) | 72 ± 10 |
| ACR, median [IQR](mg/g) | 18 [9 - 53] |
| iGFR (ml/min) | 159 ± 58 |
| Use of antihypertensives, % | 19 (43) |
| Use of renin-angiotensin system blockers, % | 19 (43) |
| Use of angiotensin receptor blockers, % | 3 (7) |
| Use of ACE inhibitors, % | 16 (36) |

Mean with SD are given for continuous variables; ACR: albumin/creatinine ratio; iGFR: iothalamate measured glomerular filtration rate. Median (IQR) for P-sclerosis.

### scRNAseq based definition of SARS-CoV-2 receptor expression in kidney tissue

To define the expression pattern of ACE2, the cell populations obtained by the scRNAseq analysis from 44 DKD and 21 LD adult human kidney tissue samples (from 18 biopsies) were clustered based on the transcription profiles of individual cells. A combined analysis of 111035 cells from the two data sets with a resolution (clustering granularity) of 0.6 defined 21 cell clusters (Figures 2A and 2B UMAP plots). Cells from both sample sources populated 18 of these, indicating the presence of cells with comparable identity in both sample sources (Supplementary Figure 1). The 18 cell clusters covered the entire spectrum of kidney cell types found along the nephron and tissue resident immune cells. In addition, we identified two DKD specific clusters (disease-specific (DS) and disease-specific thick ascending loop of Henle (DS_TAL)) and one LD specific cluster (transitional principal cell-intercalated cluster (tPC-IC)). The violin plots in Figure 2C and 2D show a restricted, cell-type specific mRNA expression of ACE2 in proximal tubular epithelial cells (PTEC), identified using standard transcript markers including cubilin (CUBN^31^). ACE2 was predominantly expressed in the CUBN expressing PTECs found in a PTEC cluster and a hybrid cluster (containing both distal limb of Loop of Henle (DTL) and PTECs) in both LD and DKD. For the remaining analysis of this study, we focused on the CUBN positive PTEC cells from those two clusters.

**Figure 2.**
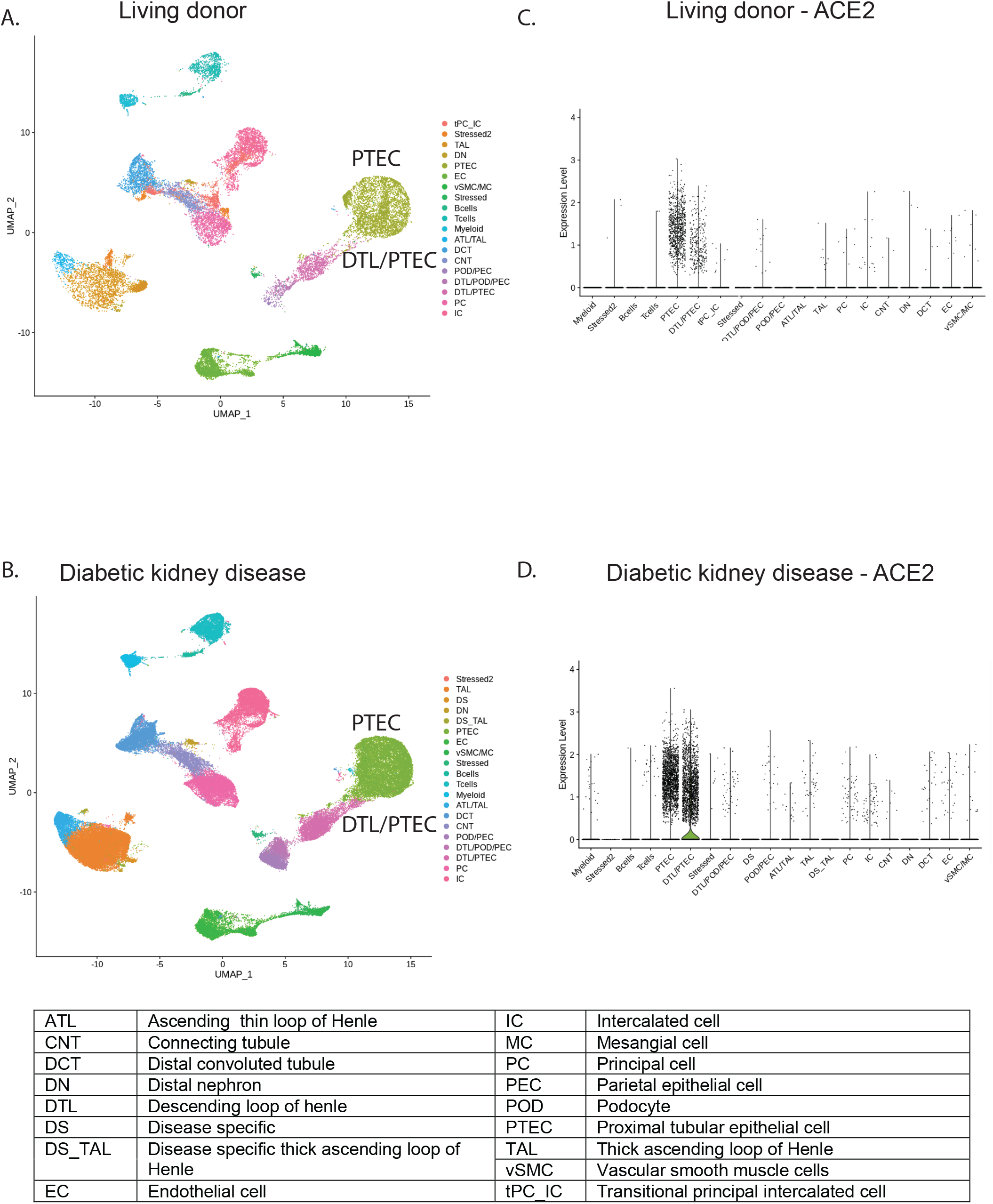
Unsupervised clustering of cells from living donor and diabetic kidney disease. UMAP plots showing the distributions in unsupervised clustering of (A) 25,163 living donor kidney cells into 19 clusters (B) 85,872 cells from DKD into 21 clusters. 22% of the total cells in both DKD and LD were CUBN positive proximal tubular epithelial cells (PTEC). The violin plots for (C) LD and (D) DKD show cell type specificity of ACE2 expression.

### Localization of ACE2 in kidneys

Cellular localization of ACE2 transcripts via *in situ* hybridization (ISH) in the kidney is shown in Figure 3 and is consistent with ACE2 expression only in proximal tubules and focally in parietal cells. Representative images from two control kidney biopsies (biopsies taken from two living donor kidneys at time of transplant) after ISH using ACE2 specific probes reveal punctate signal of ACE2 expression in PTEC, but not in distal tubular cells, glomeruli, or arterioles (Figure 3A and B). A few parietal cells also show positive signal (arrowhead in inset of Figure 3A). A clear increase in ACE2 signal in PTEC is evident in representative images from two kidney biopsies with mild features of diabetic nephropathy (Figure 3B and C) and two with advanced diabetic nephropathy (Figure 3D and E). Focally parietal cells also show increased signal (Figure 3C inset), but other cell types are again negative. Loss of signal is seen in areas of interstitial fibrosis and tubular atrophy (Figure 3E), concomitant with loss of proximal cells.

**Figure 3.**
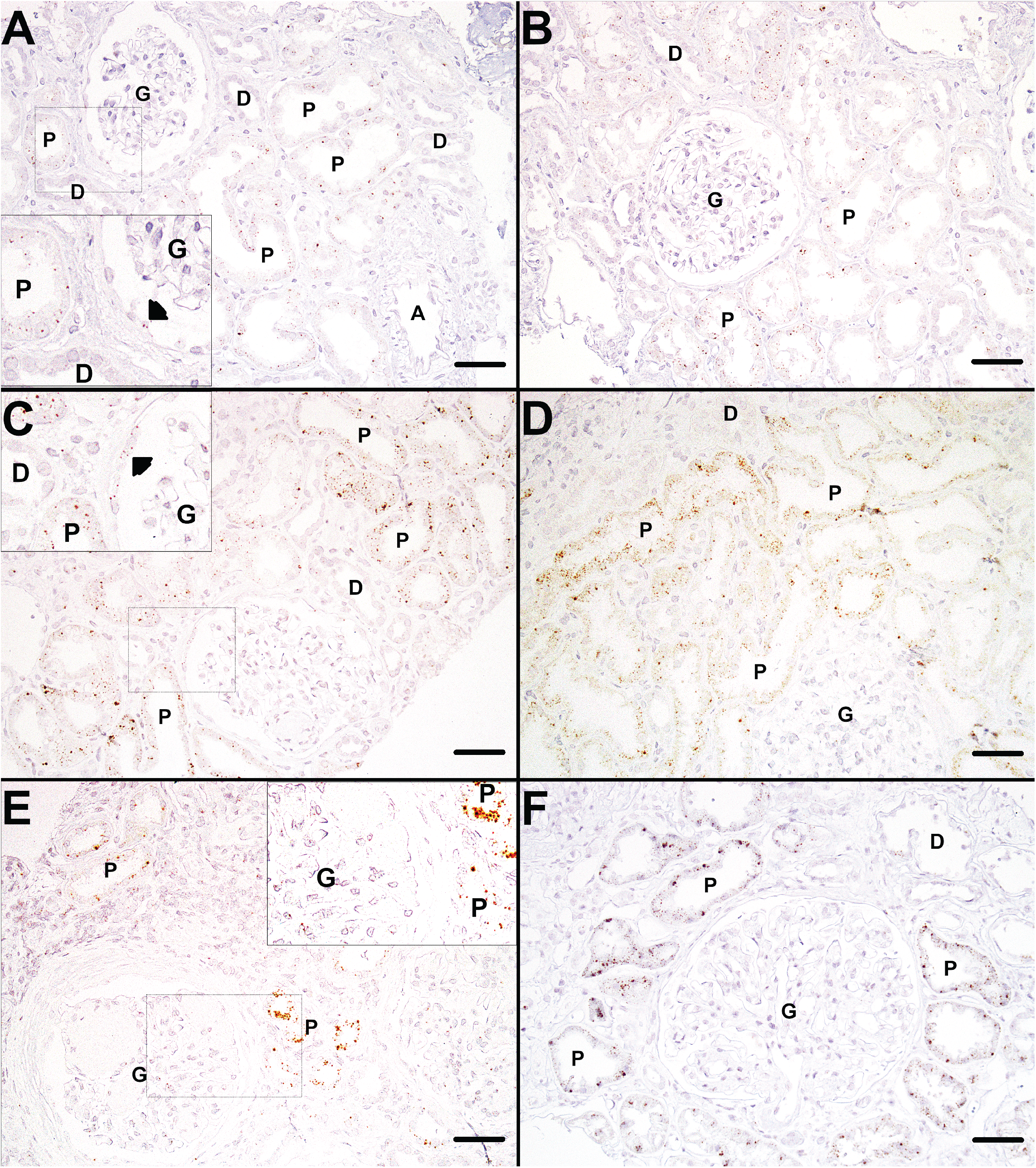
*In situ* detection of ACE2 in de-identified living donor and diabetic kidney disease tissue. *In situ* hybridization for ACE2 mRNA in two living donor reference control biopsies (A, B), two renal biopsies from patients with mild diabetic nephropathy (C, D), and two cases of advanced diabetic nephropathy (E, F). Small brown dots representing ACE2 mRNA transcripts are seen scattered at low density in proximal tubules (P) of control biopsies (A, B) and at higher density in mild (C, D) and advanced diabetic nephropathy (E, F). Small brown dots can also be seen in parietal cells in control (A) and mild diabetic nephropathy (C). No ACE2 transcript signal is seen in distal tubules, glomeruli, or arterioles. Magnification 200X. Scale bars: 50μm. Small letters denote A -arterioles; D-distal tubules; G - glomeruli; and P - proximal tubules. Arrowhead points to parietal cells.

### Defining proximal tubule ACE2 co-regulated gene programs

To define the functional context of ACE2 expression in PTEC, we first identified genes specifically expressed in PTEC with detectable expression of ACE2 (ACE2+). To this end, we first compared the expression profiles of ACE2+ to the PTEC with no detectable ACE2 expression (ACE2-) in LD and DKD, respectively. The resulting gene expression signatures (ACE2+ signatures) consisted of genes with increased expression in ACE2+ compared to ACE2-in LD (LD ACE2+ signature, Supplementary Table 2) and DKD (DKD ACE2+ signature, Supplementary Table 3).

In order to define the DKD-specific component of the ACE2+ signature, we next identified disease-induced genes, i.e., those upregulated in DKD ACE2+ compared to LD ACE2+ (DKD-induced ACE2+ signature, Supplementary Table 4). Table 2 provides a summary of the key ACE2+ gene signatures defined in this manuscript.

**Table 2:** ACE2+ signatures defined from single-cell sequencing data.

| Signature | Definition | Figure | Table |
| --- | --- | --- | --- |
| LD ACE2+ signature | Genes with increased expression in ACE2+ vs ACE2- PTEC in LD | Not shown | Supplementary Table 2 |
| DKD ACE2+ signature | Genes with increased expression in ACE2+ vs ACE2- PTEC in DKD | 4A | Supplementary Table 3 |
| DKD induced ACE2+ signature | Genes with increased expression in DKD ACE2+ PTEC compared to LD ACE2+ PTEC | Supplementary Figure 2 | Supplementary Table 4 |
| COV ACE2+ signature | Genes with increased expression in ACE2+ vs ACE2- PTEC in COV | 7A | Supplementary Table 11 |

Abbreviations: LD – living donor, DKD- diabetic kidney disease, PTEC- proximal tubular epithelial cells, COV- COVID-19 associated disease that required hospitalization

### Functional characterization of the ACE2 co-regulated gene programs in proximal tubular epithelial cells

To functionally characterize the molecular machinery expressed in ACE2 + cells in DKD, we projected the DKD ACE2+ signature genes onto the HumanBase functional network that represents biological processes and pathways active in PTEC. Intuitively, this functional network is constructed by probabilistically integrating a large compendium of thousands of public omics datasets (including expression, protein–protein interaction and motifs) to predict how likely it is that two genes act together in biological processes that function in PTEC^32, 33^. We clustered the DKD ACE2+ signature genes based on this PTEC functional network (Figure 4A, Supplementary Table 3). The resulting modules (Figure 4A, Supplementary Table 5) contained key processes of tubular function and failure, including nephron development, brush border assembly, and urate and ion transport (M7), general cellular processes such as cell cycle control (M1, M3), as well as disease signals including stress-activated signaling (M5), inflammatory pathway activation (JAK-STAT signaling, M4), and Wnt signaling (M3, M7). Importantly, we also identified key processes important to both host response and viral entry and life cycle, including viral entry (M4), viral genome replication (M1, M4), viral gene transcription (M1, M3), endomembrane organization and transport (M2, M3, M4, M7), and viral budding (M2). Immune processes were significantly enriched across multiple modules and encompassed processes including innate immune responses (M1, M4) and response to tumor necrosis factor and interferons alpha, beta, and gamma (M4), cytokine production (M4), and macrophage activation (M4).). Thus, this signature contains both key elements of documented DKD pathophysiology and processes generally associated with viral infections.^34, 35^

**Figure 4.**
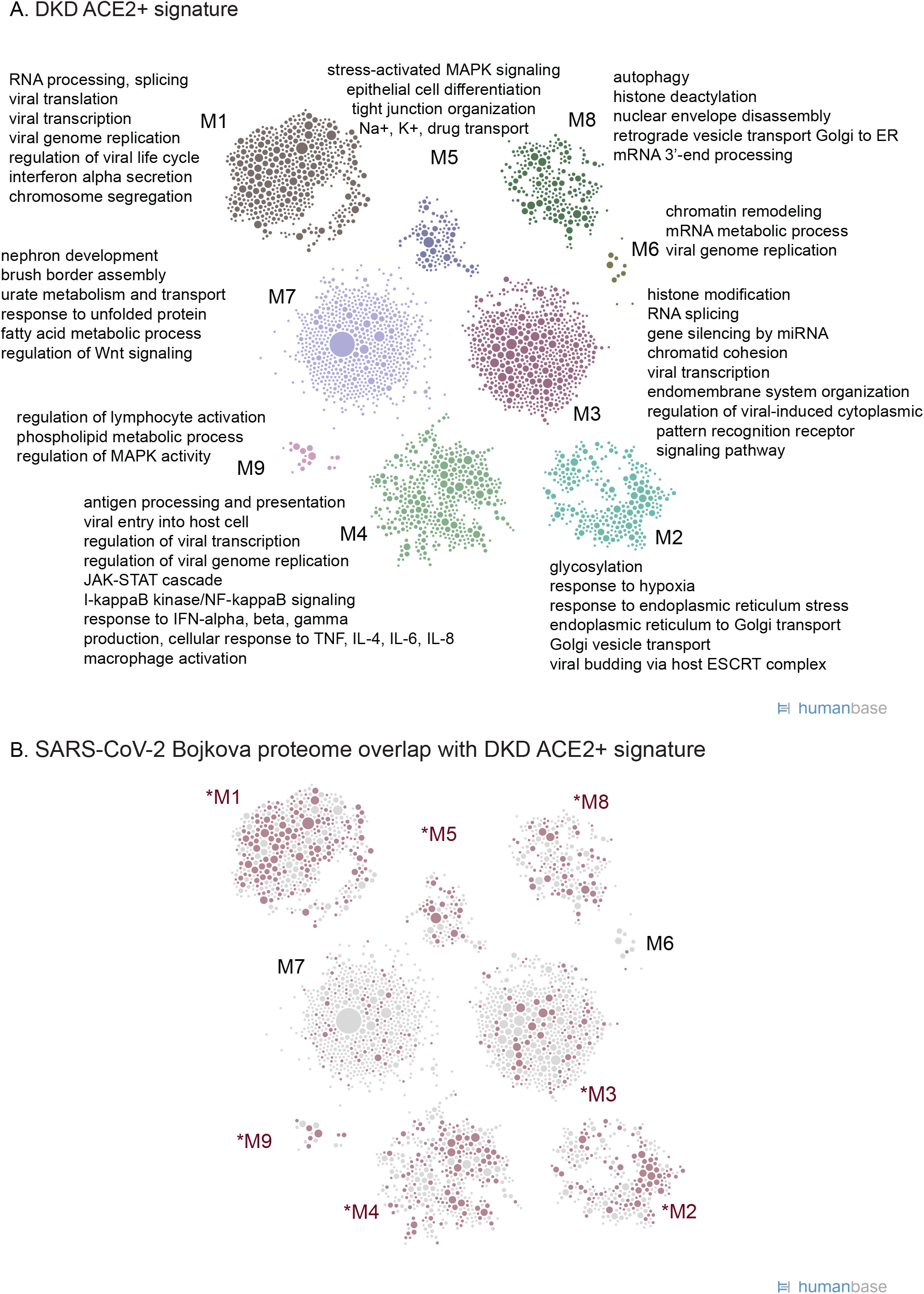
Functional summary of diabetic kidney disease ACE2+ expression signature. (A) ACE2+ co-expression signatures were used for community clustering in a PTEC-specific functional network to identify enriched processes and pathways in DKD biopsy samples. (B) Highlight of ACE2+ signature genes (red circles) shared with the set of proteins that increase expression in response to SARS-CoV-2 infection (Bojkova proteome^37^ Supplementary Table 7). Red text indicates modules with the strongest overlap (p < 0.05), highlighting modules and processes in the ACE2+ signature relevant to SARS-CoV-2 infection.

The signature above describes processes in ACE2+ compared to ACE2-PTEC in biopsies of patients with DKD. To discover which aspects of DKD could be linked to a more severe COVID-19 disease progression, we identified genes upregulated in ACE2 + DKD compared to LD (Supplementary Table 4). Analysis of these genes demonstrated prominent signals reflecting induction of processes such as inflammation and immune signaling, regulation of kidney size and morphogenesis of kidney epithelium, consistent with reported impacts of diabetic pathology on proximal tubules and PTEC (Supplementary Table 6, Supplementary Figure 2A). Notably, many genes and processes connected to RNA splicing and several aspects related to viral biology were also induced in DKD.

To explore the overlap of DKD and COVID-19 molecular programs, we examined how the DKD ACE2+ signature genes relate to those perturbed during a SARS-CoV-2 infection (Table 3). We compared the DKD ACE2+ signature genes with published SARS-CoV-2 relevant gene sets: host proteins that interact with SARS-CoV-2 proteins (Gordon Interactome^36^), proteins whose expression significantly changes in response to infection with SARS-CoV-2 (intestinal epithelial cell line Caco-2^37^), genes whose expression significantly changes in response to SARS-CoV-2 infection in primary NHBE cells, and a curated set of genes impacted by diverse coronaviruses^20^. In each comparison, we found that a significant fraction of the DKD ACE2+ signature was shared with each of the different SARS-CoV-2 gene lists (Table 3), suggesting a set of common responses to SARS-CoV-2 infection that is shared with the DKD ACE2+ signature. Specifically, a significant fraction of the DKD ACE2+ signature genes (29%, p-value < 2.2×10^-16^) also change protein expression in response to SARS-CoV-2 infection^37^, showing consistency at both levels of regulation (Figure 4B, Supplementary Table 7). We found that processes in nearly all the modules of the DKD ACE2+ signature contained genes that were specifically perturbed in SARS-CoV-2 infected cells (Supplementary Table 8). However, the module with functional enrichments most relevant to kidney biology (M7) was not enriched in SARS-CoV-2 processes, reflecting the kidney specificity of the DKD ACE2+ signature. The transcripts in the DKD-induced ACE2+ signature (Supplementary Figure 2A, Supplementary Table 4) also significantly overlapped with specific SARS-CoV-2 infection gene sets (Table 3). The shared signals are focused on translation, anti-viral responses and antigen presentation and related signaling (Supplementary Figure 2B, Supplementary Table 9). Taken together, these functional analyses suggest that expression programs active in ACE2-expressing PTEC in DKD could interact with viral infection and modulate host response.

**Table 3.** Overlap between ACE2 expression signature lists and previously reported SARS-CoV-2 associated gene sets. The size of each gene set is indicated in parentheses next to its name.

| **Number of genes**<br>**in overlap** | **DKD ACE2+**<br>**signature (3734)** | **DKD-induced**<br>**ACE2+ (839)** | **COV ACE2+**<br>**signature**<br>**(2895)** |
| --- | --- | --- | --- |
| Bojkova_proteome (2666) | 1073 | 185 | 809 |
| Gordon_interactome (332) | 123 | 4 | 84 |
| Zhou_interactome (119) | 53 | 3 | 33 |
| Blanco-Melo_NHBE (553) | 126 | 37 | 76 |
| **Fold enrichment** |  |  |  |
| Bojkova_proteome (2666) | 2.16 | 1.65 | 2.10 |
| Gordon_interactome (332) | 1.98 | 0.29 | 1.75 |
| Zhou_interactome (119) | 2.39 | 0.60 | 1.92 |
| Blanco-Melo_NHBE (553) | 1.22 | 1.59 | 0.95 |
| **p-value** |  |  |  |
| Bojkova_proteome (2666) | <2.2e-16 | 1.01e-12 | <2.2e-16 |
| Gordon_interactome (332) | 1.78e-15 | 0.9996 | 1.21e-07 |
| Zhou_interactome (119) | 7.95e-11 | 0.88 | 0.00012 |
| Blanco-Melo_NHBE (553) | 0.0079 | 0.0036 | 0.71 |

### Association of ACE2 mRNA expression in proximal tubular epithelial cells with clinical features

We analyzed the association between ACE2 gene expression and clinical measures of DKD to identify potential contributors to elevated ACE2 expression and consequent effects on disease progression. Higher expression of ACE2 was observed in CUBN positive PTEC in DKD compared to LD (Wilcoxon rank sum test p value < 0.009, log fold change = 0.05, Figure 5A). Meanwhile, proteases implicated in coronavirus infection^36, 38^, including ANPEP, BSG, CTSL, DPP4, ENPEP, FURIN and TMPRSS2, were also detected at varying levels in PTEC in LD and DKD (Figure 5B). In testing for individual-level associations with clinical characteristics, ACE2 expression levels in PTEC per participant was neither associated with baseline characteristics in the DKD cohort (e.g. age and gender, Figure 5C and D) nor treatment exposures to RAAS inhibitors (Figure 5E-G). No consistent relationships were found between ACE2 expression in PTEC and parameters of structural injury across the spectrum of DKD available to us in this cohort.

**Figure 5.**
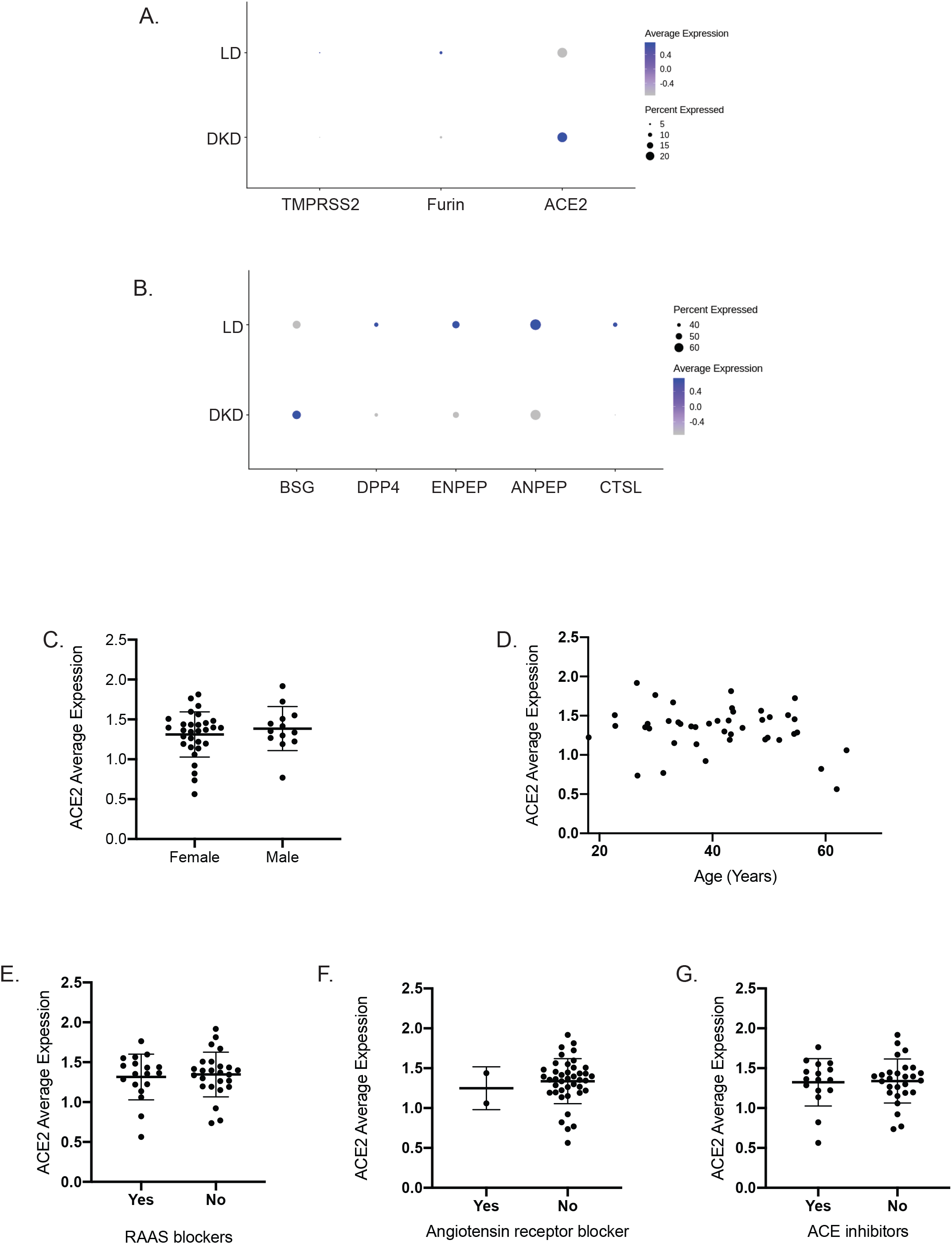
SARS-CoV-2 related host gene expression in diabetic kidney disease biopsy samples and relationships to clinical phenotypes. <u>(A and B) Expression of ACE2 and related proteases in PTEC</u> Dot plot showing the expression of ACE2 and proteases in PTEC from DKD and LD tissue analyzed by single cell analysis. Color indicates expression level and the size of the dot indicates the percentage of cells expressing the gene. Proteases were grouped into two sets based on the overall expression pattern in proximal cells (Panel A, expression in < 30% of the cells; Panel B, expression in >=30% of the cells). <u>(C-G) Clinical Associations of ACE2 expression in DKD</u>. Relationships between ACE2 expression in PTEC and (C) biological sex (D) age (E) exposure to any Renin-Angiotensin-Aldosterone-System (RAAS) Blockers; (F) exposure to Angiotensin receptor blockers; (G) exposure to Angiotensin Converting Enzyme (ACE) inhibitors were examined. No significant correlations between clinical factors and ACE2 expression were found in the DKD cohort.

### Transcriptional profile of urine-derived proximal tubular epithelial cells from COVID-19 patients

Finally, to determine the COVID-19 induced cellular responses in kidneys, we used scRNAseq to analyze urine-derived PTEC from patients hospitalized for COVID-19. The urine of thirteen COV patients yielded 25,791 cells that passed our quality control threshold. Unsupervised clustering of these cells produced seven clusters, including a PTEC/kidney cell, three immune cell types, a red blood cell, a urothelial and an undifferentiated cell cluster (Figure 6A, Supplementary Figure 3). Of the cells in the PTEC cluster 13% expressed ACE2. In addition to ACE2, proteases including ANPEP, BSG, CTSL, DPP4, ENPEP, FURIN and TMPRSS2 were also expressed in this cluster (Figure 6B). Supplementary Table 10 provides a list of the PTEC markers identified in COV. Functional network analysis of the COV ACE2+ signature identified ten modules, with modules M3, M5, and M6 enriched for viral processes, including viral genome replication and viral gene expression, consistent with the patients’ COVID-19 state (Figure 7A, Supplementary Table 12). We also identified a module (M8) enriched in functions that reflect PTEC processes, such as brush border assembly, ion transport, and urate metabolic process. Other enriched processes include cellular functions critical for SARS-CoV-2 viral infection, such as translation, endomembrane system remodeling and RNA metabolism pathways (M1, M3, M5, M7). Modules enriched in mitochondrial pathways, autophagy processes indicative of cell stress were also present in the signature (M2, M7).

**Figure 6.**
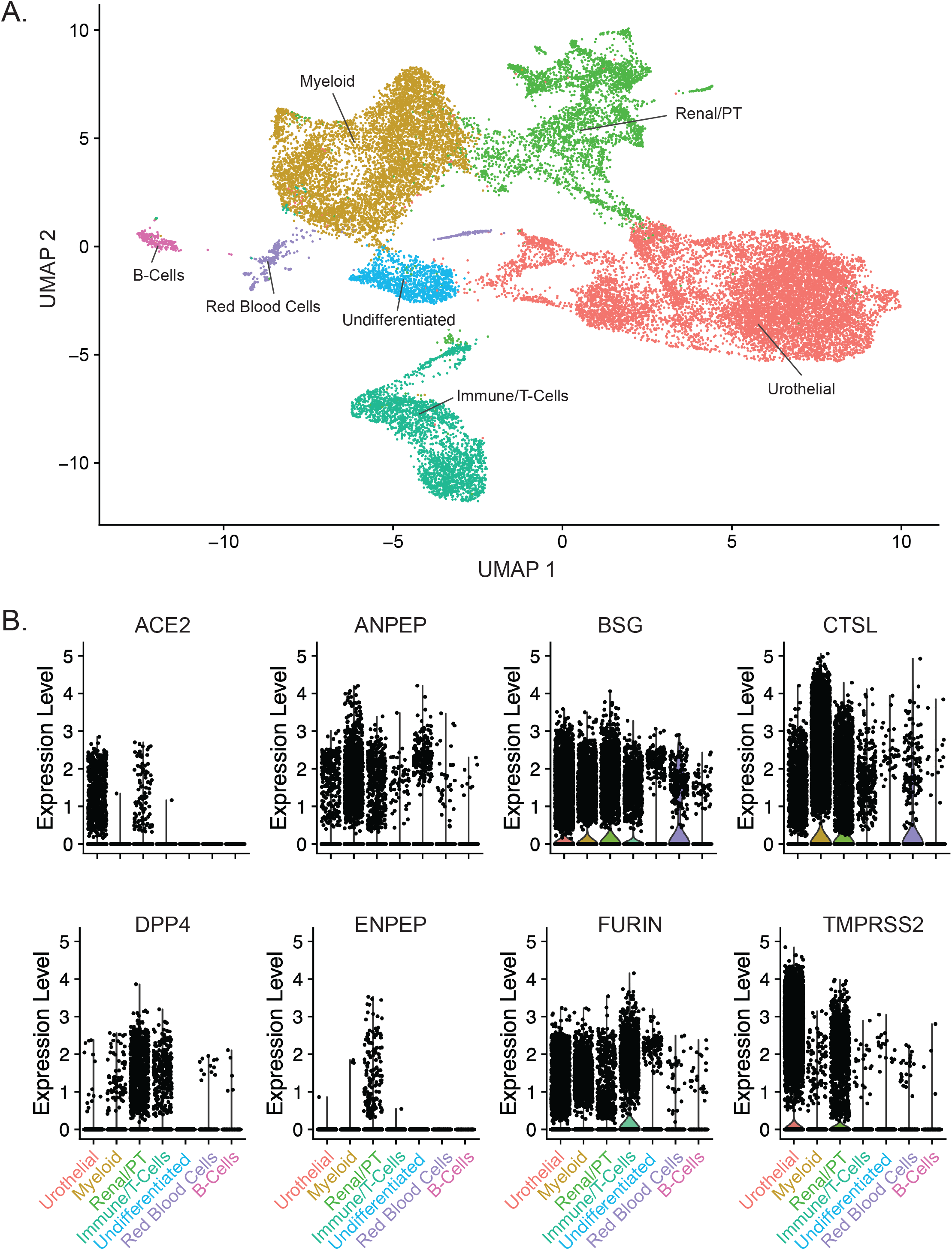
Single cell analysis of cells isolated from the urine of patients hospitalized with COVID-19 (COV) (A) UMAP plot showing the seven clusters identified from 25,791 cells. (B) Violin plots of SARS-CoV-2 associated proteases, ACE2, ANPEP, BSG, CTSL, DPP4, ENPEP, FURIN and TMPRSS2 in the seven identified cell types.

Overall, the COV ACE2+ signature significantly overlaps the DKD ACE2+ signature (p-value < 2.22×10-16) and is functionally consonant with processes critical for viral infection and immune response (Figure 7B, Supplementary Table 13). Interferon secretion and response pathways are also represented, however, a more extensive set of interferon receptors, interferon stimulated genes and cytokines are differentially expressed in the COV ACE2+ signature (Supplementary Table 14). Most aspects of the COV ACE2+ signature overlap significantly with the DKD ACE2+ signature, with the notable exception of a module enriched in mitochondrial pathways (M2) specific to the COV ACE2+ signature, which may reflect a stress signal in kidney cells shed in the urine (Supplementary Table 15). We then compared the COV ACE2+ signature to other reported SARS-CoV-2 datasets^20, 36, 37, 39^ and found significant concordance (Table 3) with key viral processes such as vesicle organization, regulation of viral processes, interferon response, stress signaling, and endomembrane organization and transport (Figure 7C, Supplementary Table 16).

**Figure 7.**
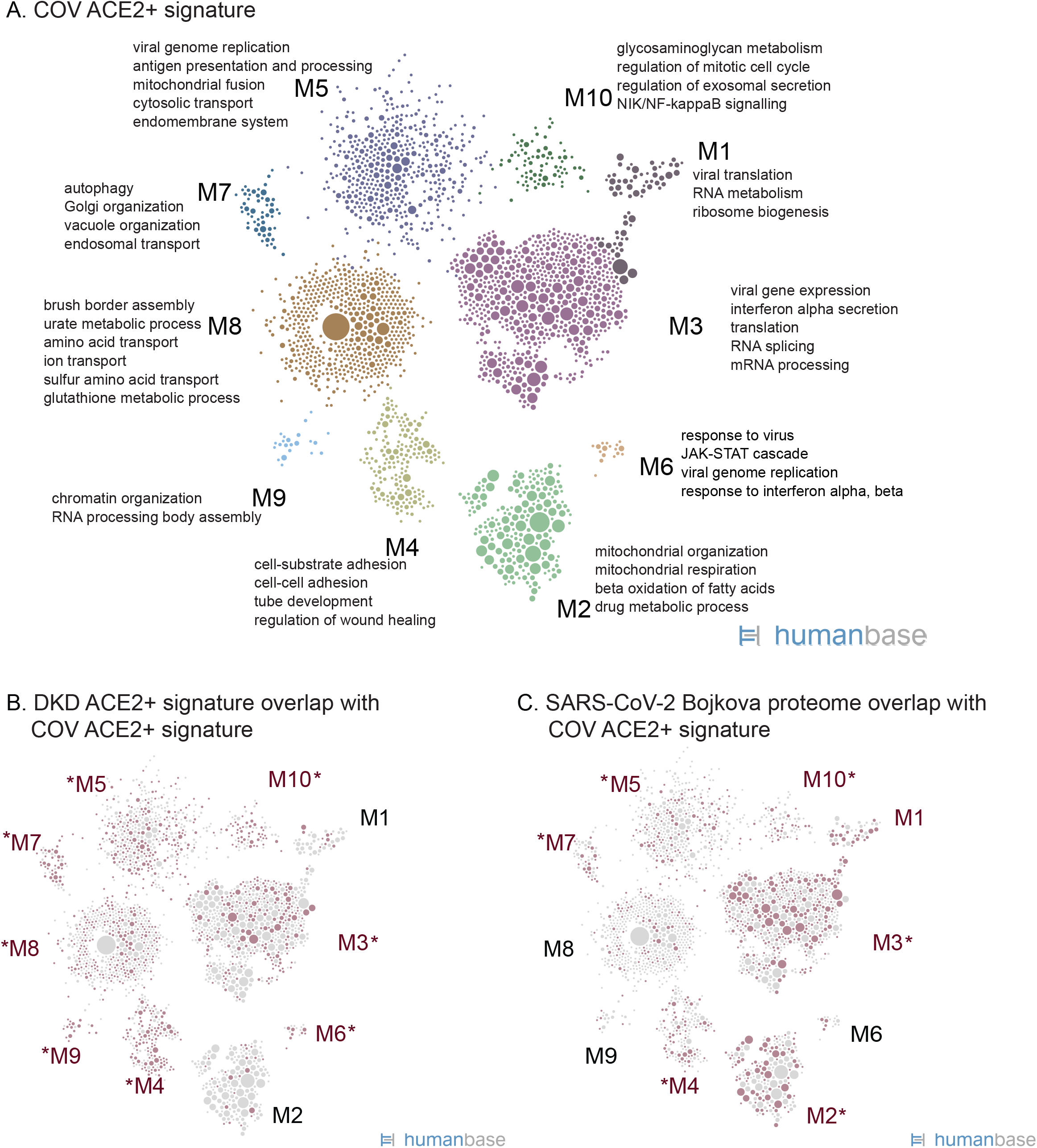
Functional summary of COV ACE2+ expression signature. (A) Community clustering and enriched processes of the ACE2+ expression signature in PTEC isolated from urine of patients with COV. (B) Highlight of genes (red circles) from the DKD ACE2+ signature (Figure 4A, Supplemental Table 3) that overlap the ACE2+ signature in PTEC from COVID-19 patients. Red text indicates modules with strongest gene overlap (p < 0.0105), thus indicating ACE2+ associated processes shared between PTEC in diabetic kidneys and PTEC in the context of COVID-19. (C) Highlight of ACE2+ signature genes (red circles) shared with the set of proteins that increase expression in response to SARS-CoV-2 infection (Bojkova proteome^37^, Supplementary Table 7).

## Discussion

With the pandemic spread of SARS-CoV-2 and the increased morbidity and mortality of COVID-19-associated disease in patients with diabetes and kidney disease, it is imperative to define the underlying mechanism of the excessive risk in patients with diabetes and rapidly develop protective strategies for risk reduction. Autopsy studies of patients with COVID-19 support kidney tropism of SARS-CoV-2 and that even within the kidney^10, 15, 38^, expression of molecules like ACE2, that mediate viral entry may be cell type-specific^15^. Susceptibility and infection of *ex-vivo* kidney cultures are consistent with the notion of a direct SARS-CoV-2 infection of the kidney tissue with viral tropism being limited to specific cell types^10, 15, 38^. ACE2 is an enzyme that governs several pathways in kidney cells, including RAAS, that are critical to kidney function. Therefore, we sought to determine whether the gene expression signature of ACE2-expressing PTEC in kidneys was different from ACE2 negative PTEC and whether this signatures were regulated between DKD and LD PTECs with the ultimate aim of finding markers of viral susceptibility.

We first compared the expression of ACE2 and the other reported SARS-CoV-2 entry factors in healthy individuals and in patients with DKD. ACE2 intra-renal gene expression was localized predominately to PTEC in surveillance kidney biopsies obtained from LD and in research kidney biopsies from patients with DKD. The only other nephron segment with low ACE2 level expression signals were parietal glomerular epithelial cells, consistent with their shared developmental origin. In contrast to published murine data^40-45^, no significant expression was seen in glomerular podocytes or in the different endothelial cell compartments evaluated in the scRNAseq profiles. Consistent with our studies of expression and localization of ACE2, PTEC appear to be the main cell type of SARS-CoV-2 infection in the kidney^10, 12, 15^ As opposed to findings in other organ systems, little co-expression of ACE2 with the TMPRSS2 co-receptor for SARS-CoV-2 viral entry was observed, but robust expression of other cell surface proteases including cathepsin-L, which have been associated with viral infection and uptake, were detected ^15, 38^.

Even though none of the DKD patients in this study were infected with SARS-CoV-2 at the time of biopsy, our pathway analysis showed that processes including viral infection, protein processing and antigen presentation were enriched in ACE2 expressing PTEC in DKD. ACE2 is co-expressed in PTEC with a set of genes that also function in establishment of viral replication, host responses, and innate immunity. These results, taken together with the consistent functional themes observed between DKD and COV ACE2+ signatures, suggest that the gene sets which are co-expressed in PTEC with ACE2 in DKD may establish a cellular program that specifically interacts, in these cells, with processes induced by viral infection and host immune response. The ACE2 associated pathways could interact in two ways with SARS-CoV2 infections of PTEC. As pathways associated with viral infection are already upregulated in diabetes patients, this could explain the higher susceptibility of this patient population. If viral infection of ACE2+ PTEC activates pathways that are already upregulated in diabetes, this cumulative and exacerbating activation might lead to kidney damage.

As in the pulmonary system^46, 47^, it can be expected that ACE2 expression renders PTEC vulnerable to initial SARS-CoV-2 viral entry and that the ACE2+ co-expressed genes define a program ready to engage the incoming virus. Ex vivo studies in kidney organoids, indeed, support the ACE2 dependence of SARS-CoV-2 infection of PTECs^48^ . This program includes genes that may promote successful viral infection, and subsequent increased viral-induced cytopathic effect in DKD, especially as viral replication and processing machinery are upregulated in the ACE2+ PTEC in DKD (Figure 4A). Signature genes reported to change expression in response to SARS-CoV-2 infection (highlighted in Figure 4C and listed in Supplementary Table 7 “Bojkova proteome List”) are genes that may directly interface with a SARS-CoV-2 infection in kidney PTEC. To understand the processes impacted by these genes and develop hypotheses for experimental exploration, we clustered the PTEC functional network with this overlap gene list and identified enriched pathways (Supplementary Table 17). Genes involved in endocytosis and endomembrane transport (DNAJC13, CHMP3, SNX9, PDCD6IP) and membrane organization (GOLGA4, ANXA2, ANK3) could facilitate viral entry, replication and exit^49^, and proteins that interact with viral gene transcription (DHX9, HDAC1, NUCKS1, SMARCA4, SMARCB1, CHD1, and RSF1), viral RNA translation (EIF3A, MCTS1, SSB, and DHX9) and viral genome replication (DDX3X, PPIE, and PABPC1) could promote productive replication. Inflammation and interferon response genes included in this overlap gene set include IFNGR1, ILF3, TNFAIP2, TNFAIP8, and successful host defense of SARS-CoV-2 may hinge on their activity. It is intriguing that a number of the ACE2+ DKD signature genes encode for proteins that interact or have been predicted to interact with SARS-CoV-2 proteins^36^, and are involved in anti-host defense response; including the membrane “M” protein (suppresses type 1 interferon responses and modulate cytokine responses)^50, 51^, the nucleoprotein “N” (inhibits autophagy and apoptosis of infected cells through mTOR)^47^, Orf3a (may modulate IL-1B)^52, 53^, and Nsp6 (inhibits phagosome expansion and targeting of viral products to lysosome)^54^. These interactions may simply reflect an element of the functional host response program to cellular stress in DKD. If these physical interactions occur during the course of SARS-CoV-2 infection and involve negative functional regulation of host-defense proteins, ACE2+ PTEC could be impaired in their ability to mount an appropriate host response, providing one explanation of increased susceptibility to SARS-CoV-2.

The inflammatory signals observed in the DKD PTEC are expected in the context of chronic kidney disease as reported previously by studies in this and other DKD patient populations^55, 56^ However, these data could also indicate that this inflammatory state could be maladaptive for a response to viral infection. Recent work on lung epithelial cells infected with SARS-CoV-2 has shown a blunted interferon response despite robust cytokine production still being present^39^; a potential hallmark of SARS-CoV-2 pathology. If a comparably asymmetric inflammatory response is already present in DKD kidney tissue, viral infection could amplify the pre-existing inappropriate stress response, increasing cytopathology and enabling further viral propagation. The PTEC shed in the urine of COVID-19 patients show an interferon activation and response signature, indicating that interferon response to SARS-CoV-2 infection can be robust in the kidney. The cellular inflammatory state of ACE2+ cells may permit enhanced viral pathogenesis and replication in multiple organ systems, and this linkage could provide an evolutionary driving factor to select ACE2 for viral entry.

The networks observed in genes upregulated in ACE2-positive versus -negative PTEC in DKD and COV allow for a wide range of exploratory studies to generate starting points for functional evaluation. Given the urgent need for novel therapeutic options, intersecting the ACE2 regulatory networks with the complex landscape of interactions between drugs and the gene signature defined here can help to identify therapeutic targets for further study. We identified ACE2+ co-regulated genes annotated in DrugBank as targets of medications regularly taken by patients with DKD. For example, the insulin receptor (INSR, in M4 in Figure 4A, Supplementary Table 3) and the IGF-1 receptor (IGF1R, in M4 in Figure 4A) are overexpressed in ACE2+ PTEC as is DPP4 (in M7 in Figure 4A), a protease associated with entry of other coronaviruses and a key factor in glucose metabolism, targeted by DPP4 inhibitors already in clinical use. These interactions suggest that the ACE2+ co-expression signature may be influenced by a number of commonly used drugs and can form the starting points for further mechanistic and epidemiological studies.

As data emerged on the role of ACE2 as a receptor for SARS-CoV-2, significant concerns were raised about risks of RAAS inhibitors frequently prescribed in patients with diabetes and chronic kidney diseases. Based on animal models RAAS inhibitors were predicted to increase ACE2 levels raising the potential concern for higher COVID-19 morbidity and mortality^29, 57, 58^ However, our findings in the DKD cohort do not support the animal model data and do not provide any evidence associating ACE2 expression in PTEC to RAAS inhibition. In parallel, a series of case control, database and Electronic Health Record studies did not find an association of RAAS inhibitors with poor outcomes in three independent cohorts of patients with COVID-19 and RAAS exposures^59-61^, consistent with our scRNAseq studies.

ACE2 gene expression studies of kidney tissue to date have been inconclusive concerning regulation in kidney disease ^42, 62-64^ A strength of our study is the definition of the ACE2 expression to the cellular context of PTEC in kidney disease and in patients with COVID-19, with the ACE2 expression in PTEC consistent with published work from reference kidney tissue^12, 65, 66^. For the study of gene expression in kidney disease, the resolution of single cell studies is critical because disease-associated loss of PTEC cells confounds bulk tissue analysis of ACE2 expression. The PTEC dominant ACE2 scRNAseq expression data presented here match the pattern observed in the human protein atlas^67^ (https://www.proteinatlas.org/, Supplementary Figure 4).

Our study is also limited by assessing mRNA levels, which only capture one of several levels of regulation of ACE2 function. For this reason, our analyses focused on the changes in transcriptional programs as a functional readout and not protein activity directly. However, as noted above, our mRNA findings for ACE2 are consistent with protein expression specificity from the Human Protein Atlas. In addition, the ACE2 signatures and reported SARS-CoV-2 infection gene sets do correspond at both mRNA and protein levels. We focused our analysis on ACE2 as the best characterized SARS-CoV-2 receptor. As additional receptors and mechanisms of SARS-CoV-2 cellular entry might emerge, our data sets can be mined for regulation and co-expression in a disease-specific context.

The hypotheses proposed above on the potential interactions between the molecular programs co-expressed with ACE2 in PTEC can form the starting points for experimental validation as critical next steps. Experimental model systems of kidney SARS-CoV-2 toxicity are currently under development, and intriguing data on human kidney organoid systems show ACE2 expression in the tubular cell cluster. ACE2, in the correct functional context, enables SARS-CoV-2 virus to infect tubular epithelial cells and a therapeutic strategy involving soluble ACE2 reduced cytotoxicity in the organoid system ^48, 68^, a strategy with soluble ACE2 has already been tested in acute respiratory distress syndrome and is available for clinical testing. Combining the organoid model system with the human interaction maps generated in this study can form an experimental framework for mechanistic studies testing the reported associations for their functional impact.

In summary, our work identifies the regulation and associated cellular machinery of ACE2 and associated SARS-CoV-2 co-receptors in PTEC in kidney health, metabolic and viral disease. The SARS-CoV-2 receptor associated networks are now available (hb.flatironinstitute.org/covid-kidney) to seed further research into urgently needed therapeutic strategies for COVID-19.

## Methods

### Study population

#### DKD biopsy cohort (DKD)

An early DKD cohort was selected as the discovery cohort given the impact of COVID-19 on individuals with diabetes and kidney disease^3, 5, 8, 14, 29, 30^. Between 2013 and 2017, 141 American Indians (39 men, 102 women) from southern Arizona with type 2 diabetes were enrolled in an observational study of DKD that included annual research examinations. Glomerular filtration rate (GFR) was measured at each of these examinations by the urinary clearance of iothalamate^69^. Each participant also underwent a research kidney biopsy used for quantitative morphometry and for compartment-specific tissue expression studies. For kidney biopsies obtained from the last 44 participants enrolled between 2016 and 2017, processing of the kidney specimens was modified to enable scRNAseq studies using kidney biopsy tissues. These 44 participants (DKD) were included in the present study.

At each annual research examination, medicines taken by the participants were recorded. Blood pressure (BP) was measured with the participant resting in a seated position, and height and weight were obtained with the participant dressed in light clothing without shoes. Body mass index was defined as weight in kilograms divided by the square of height in meters. Urine and serum creatinine were measured by an enzymatic method. Urinary albumin excretion was assessed by nephelometric immunoassay and expressed as the albumin/creatinine ratio (ACR) in mg/g creatinine. GFR and HbA1c were measured by high performance liquid chromatography. Because of the level of obesity in the participants, absolute GFR (ml/min) was reported, uncorrected for body surface area^70^. Clinical and demographic features and the antihypertensive medicines taken by the DKD cohort at the time of the kidney biopsy are shown in Table 1. Participants had mean diabetes duration of 12.2 ± 7.5 years, HbA1c of 9.2 ± 2.4 %, iGFR of 159 ± 58 ml/min with one patient GFR <60ml/min, and median ACR of 18 [9-53] mg/g (25 persons had a normal ACR <30 mg/g, 14 had moderate albuminuria of 30-300 mg/g and 3 had severe albuminuria >300mg/g of ACR at biopsy).

#### Allograft kidney transplant surveillance cohort (LD)

The University of Michigan has a kidney transplant surveillance program in which biopsies are performed for clinical care at implantation and 3-, 6- and 12-months post transplantation. A subset of patients in this surveillance program participate in the Human Kidney Transplant Transcriptomic Atlas study, which procures additional biopsy cores for research. Tissue samples from these biopsies were used in the present study. Reference healthy tissue 18 LD biopsies were obtained prior to perfusion and before placement in the recipient. Participant mean age was 45.1 ± 10.2 years, range (30-66), 11 of the 18 donors were female (61%), mean spot urine protein to creatinine ratio at the time of evaluation was 0.08 ± 0.04 g/g with range (0.03-0.20), mean iothalamate GFR was 100.6 ± 16.9 ml/min/1.73 m^2^ with range (81-144). Fifteen of 17 donors were white, one of Hispanic descent and one of African ancestry. All donors were nondiabetic and non-hypertensive. Technical replicates were performed on two LD biopsies with 2 and 3 biopsy segments processed in parallel.

#### COVID-19 cohort (COV)

In response to the COVID-19 pandemic at the University of Michigan Medical Center a protocol to capture urine samples from SARS-CoV-2 patients was established to allow cell and molecular biology studies of the kidney-related manifestations in COVID-19. Patients are offered participation in the study upon admission to the hospital and throughout their hospital course. 50 ml of a spot random urine sample was procured specifically for the study, immediately transported to the research laboratory on ice and processed for cell and molecular studies and participants are prospectively followed thereafter.

Patients were eligible to participate if they were adults admitted to the hospital with a positive PCR test for SARS-CoV-22 and had symptoms attributable to SARS-CoV-2. Exclusion criteria included a history of ileal conduit, bladder cancer, anuria or if the initial positive SARS-CoV-2 test was more than a month prior to urine collection. Clinical data were abstracted from the participants’ charts and summarized in Supplementary Table 1. Average participant age was 50 (SD of 17.3) and 54% were male. The median time since the first positive COVID-19 test was 11 days and 62% of the participants had AKI by KDIGO serum creatinine criteria. At the time of urine sample collection, 38% of participants had a down-trending serum creatinine and 85% required ICU admission at some point during their hospitalization. 54% of the participants had diabetes mellitus (DM).

### Kidney biopsy sample processing for single cell

Single cell transcriptomes were generated with 2-3 mg of the biopsy core samples from 44 CryoStor® (Stemcell Technologies) preserved DKD and LD biopsies. Tissue processing and single cell isolation were performed following our published protocol ^71^. Briefly, the tissue samples were quickly thawed, washed with culture medium and dissociated to single cells using Liberase™ TL (research grade, Roche) for 12 minutes at 37°C. Single cell samples were immediately transferred to the University of Michigan Advanced Genomics Core facility for further processing.

### Urine cell preparation for scRNAseq in patients with COVID-19

Urine single cell preparation followed the protocol by Arazi et al. with various modifications^72^. Briefly, COV urine was filtered through a 30 μm strainer into 50 ml tubes and centrifuged at 200 × *g* at 4°C for 10 minutes. After removing and storing the supernatant the cell pellet was washed once with 1 ml cold X-VIVO™10 medium (Cat#: 04-743Q, Lonza) and centrifuged in 1.5 ml tubes for 5 minutes at 200 x *g* at 4°C. The resulting cell pellet was then suspended in 50 μl DMEM/F12 medium supplemented with HEPES and 10% (v/v) FBS and about 50,000 cells were loaded onto the single cell droplet-based RNAseq platform. Next generation sequencing was carried out on a NovaSeq6000 (Illumina) machine generating a 200 million read depth per sample. All procedures were performed following biosafety precautions as directed by the Centers for Disease Control and Prevention and the University of Michigan guidelines.

### scRNAseq data generation and analysis

Individual cell barcoding, reverse RNA transcription, library generation and single cell sequencing using Illumina were all performed using the 10X Genomics protocol^71^. The output from the sequencer was first processed by CellRanger, the proprietary 10X Chromium single cell gene expression analysis software (https://support.10xgenomics.com/single-cell-gene-expression/software/pipelines/latest/what-is-cell-ranger). Data analyses were performed on the CellRanger output data files using the Seurat 3 R package (https://cran.r-project.org/web/packages/Seurat/index.html). As a quality control step, cells with less than 500 or greater than 5000 genes per cell were filtered out, with the lower cut off as a threshold for cell viability and the higher cut off to remove cell duplets. This study used a cutoff of <50% mitochondrial reads per cell as an additional threshold for cell viability. A combined analysis of the single cell datasets generated from the different sample sources (LD and DKD) was performed using Seurat (v3) that included the following steps: default normalization, scaling based on sample mRNA count and mitochondrial RNA content, dimensionality reduction PCA (Principal component analysis) and UMAP (Uniform Manifold Approximation and Projection), sample integration using Harmony algorithm, standard unsupervised clustering, and the discovery of differentially expressed cell type specific markers^73^. According to Seurat, the normalization step uses a global-scaling method that normalizes the gene expression measurement for each cell by the total expression, multiplies this by 10,000 (default) and log-transforms the scaled expression values. For cell type annotation we used publicly available resources including published literature, Kidney Interactive Transcriptome (http://humphreyslab.com/SingleCell/), Human Protein Atlas (HPA) (https://www.proteinatlas.org), the Epithelial Systems Biology Laboratory (ESBL) (https://hpcwebapps.cit.nih.gov/ESBL/Database/), scRNAseq data and Immgen (https://www.immgen.org/). The same analysis processes were used for the COV samples except that no Harmony-based integration step was performed.

Further functional analyses were focused on PTEC expressing detectable ACE2 (ACE2+) and without ACE2 (ACE2-) mRNA in the different samples studied. In order to identify PTEC, for the DKD analysis, we filtered for cells in the PTEC and DTL/PTEC clusters with CUBN expression, and for COV, cells with CUBN, GATM, or LRP2 expression^71, 74^.

### In situ hybridization of ACE2 in control and diabetic kidney biopsies

In situ detection of ACE2 (RNAscope® Probe Hs-ACE2, Advanced Cell Diagnostics [ACD], catalog #848151), mRNA transcripts using the RNAscope kit (ACD) was performed according to the manufacturer’s protocol by kidney pathologists able to identify cell types in kidney tissue. Sections (3 μm) sections of de-identified human kidney tissue were cut from formalin-fixed, paraffin-embedded (FFPE) blocks supplied by the University of Michigan Tissue Procurement Service. Housekeeping gene ubiquitin C (UBC, RNAscope® Positive Control Probe Hs-UBC, ACD, Catalog #310041) was used as an internal mRNA control and the bacteria DapB (RNAscope® Negative Control Probe- DapB, ACD, Catalog #310043) as a negative control gene. A horseradish peroxidase–based signal amplification system (RNAscope 2.0 HD Detection Kit-Brown, ACD, catalog 310035) was used for hybridization to the target probes, followed by color development with DAB, and the slides were counter-stained with hematoxylin (RICCA Chemical Company, catalog 3535-16). Positive staining was determined by brown punctate dots in the cytoplasm.

### Identification of differentially expressed ACE2+ co-regulated gene signatures

Differentially expressed genes were identified between ACE2+ and ACE2-PTEC in DKD, and COV samples using the FindAllMarkers Seurat function. For LD, DKD and DKD-induced signatures, all genes with Bonferroni adjusted p-value < 0.05, positive log fold change, and found in at least 10% of DKD ACE2+ PTEC cells were selected. For the COV ACE2+ signature, all genes with nominal unadjusted p-value < 0.05, positive log fold change, and found in at least 10% of ACE2+ PTEC cells were selected. (Supplementary Tables 2-4, 11).

### Overlap of differentially expressed ACE2+ co-regulated gene signatures with SARS-CoV-2 relevant gene sets

SARS-CoV-2 relevant gene sets were compiled from multiple published sources: 1) Bojkova_proteome is a list of differentially expressed proteins in Caco-2 cell line following SARS-Cov-2 infection (p < 0.05 at any time point)^37^ 2) Gordon_interactome is a set of host proteins identified as physically interacting with SARS-CoV-2 viral proteins in HEK-293T cells^36^; 3) Zhou_interactome is a literature-curated list of genes related to diverse coronaviruses^20^; 4) Blanco-Melo_NHBE is a list of differentially regulated genes in response to SARS-CoV-2 infection in normal human bronchial epithelial cells^39^. ACE2 was removed from ACE2+ co-expressed gene sets before computing overlaps. P-values were computed using the hypergeometric test with a count of 20,000 genes used as background. Specifically, p-values were computed using the R function 1 - phyper(s - 1, g1, N - g1, g2), where s is the number of genes shared between the two gene sets, N is the number of background genes, g1 is the number of genes in gene set 1, and g2 is the number of genes in the second gene set. Fold enrichment was computed as (s/g1)/(g2/N).

### Functional network analysis

To determine the biological processes and pathways in the ACE2+ differentially expressed gene sets, we performed functional network clustering in the PTEC gene functional network derived from GIANT 2.0^32, 33^ This network was generated through regularized Bayesian integration of 61,400 publicly available expression, physical interaction and other omics experiments to generate a fully connected weighted graph representing functional relationships in biological pathways in PTEC. Community clustering in the network was performed to identify tightly connected sets of genes using HumanBase.io module detection function^75^ (hb.flatironinstitute.org/covid-kidney). The network was clustered with each set of differentially expressed genes constituting each ACE2+ signature: LD, DKD, DKD-induced, and COV (Supplementary Table 2-4, 11). Specifically, for DKD and DKD-induced signatures, we clustered genes with Bonferroni adjusted p-value < 0.05, positive log fold-change, and found in at least 10% of cells. In order to cluster gene sets of similar sizes we limited the DKD signature (with a total size of 3735 genes) to the top 3000 genes by p-value significance for clustering. Due to the comparatively small number of PTEC in COV samples, ACE2+ co-regulated genes that passed a nominal uncorrected p-value threshold of 0.05, positive log fold-change, and were found in at least 10% of cells were selected. (2896 genes were clustered in the network). The Gene Ontology enrichment outputs are Supplementary Tables 5,6,12,14,16,17 and can be interactively explored at hb.flatironinstitute.org/covid-kidney.

### Statistical analysis of clinical associations

Spearman correlation was used to evaluate the association of the steady state average PTEC specific gene expression levels of ACE2 and age at time of biopsy for the 44 DKD samples. The Mann-Whitney non parametric test was applied to evaluate the gender and treatment difference of ACE2 expression in the DKD cohort.

### Data access

LD single cell data sets are searchable at NephroCell.miktmc.org. Owing to ethical considerations, privacy protection concerns, and to avoid identifying individual study participants in vulnerable populations, the Institutional Review Board of the National Institute of Diabetes and Digestive and Kidney Diseases has stipulated that individual-level gene expression and genotype data from the American Indian DKD study cannot be made publicly available. A dynamic user-friendly interface at HumanBase (hb.flatironinstitute.org/covid-kidney) is available for researchers to explore the functional networks of gene expression signatures.

### Study approval

All studies of the DKD population were approved by the Institutional Review Board of the National Institute of Diabetes and Digestive under protocol 13-DK-N151. Transplant biopsies were obtained after review and approval of the T ransplant T ranscriptomic Atlas study under HUM00150968; DKD kidney biopsies gene expression analysis was performed under HUM00002468; COVID-19 urine samples were obtained after review and approval under HUM00004729, all reviewed by the University of Michigan IRBMED.

## Data Availability

LD single cell data sets are searchable at NephroCell.miktmc.org. Owing to ethical considerations, privacy protection concerns, and to avoid identifying individual study participants in vulnerable populations, the Institutional Review Board of the National Institute of Diabetes and Digestive and Kidney Diseases has stipulated that individual-level gene expression and genotype data from the American Indian DKD study cannot be made publicly available.
A dynamic user-friendly interface at HumanBase (hb.flatironinstitute.org/covid-kidney) is available for researchers to explore the functional networks of gene expression signatures.

http://nephrocell.miktmc.org/

https://hb.flatironinstitute.org/covid-kidney

## Acknowledgements

Acknowledgements:

We thank Ms. Lois Jones, RN, Mr. Enrique Diaz, RN, Ms. Bernadine Waseta, and Ms. Camille Waseta for performing the studies in the diabetes cohort and the University of Michigan Advanced Genomics Core for providing expert technical assistance with single cell processing and sequencing. We also thank Dr. Carmen Mirabelli for reviewing this manuscript and providing valuable feedback.

This work was supported in part by the Intramural Research Program at the National Institute of Diabetes and Digestive and Kidney Diseases (DK069062) to HCL and RGN and (DK083912, DK082841, DK020572, DK092926) to RGK, by the extramural research program of the National Institute of Diabetes and Digestive and Kidney Diseases R24 DK082841 ‘Integrated Systems Biology Approach to Diabetic Microvascular Complications’ and P30 DK081943 ‘University of Michigan O’Brien Kidney Translational Core Center’ to MK, via ‘Kidney Precision Medicine Project’ *(KPMP, funded by the following grants from the NIDDK: U2C DK114886, UH3DK114861, UH3DK114866, UH3DK114870, UH3DK114908, UH3DK114915, UH3DK114926, UH3DK114907, UH3DK114920, UH3DK114923, UH3DK114933, and UH3DK114937, with* U2C DK114886 and UH3 DK114907) to MK and OGT, via the Chan Zuckerberg Initiative ‘Human Cell Atlas Kidney Seed Network’ to MK and OGT and by JDRF 5-COE-2019-861-S-B ‘JDRF and M-Diabetes Center of Excellence at the University of Michigan’ to MK. The content is solely the responsibility of the authors and does not necessarily represent the official views of the National Institutes of Health.

## For the Kidney Precision Medicine Project

*American Association of Kidney Patients, Tampa, FL:* Richard Knight *Beth Israel Deaconess, Boston, MA:* Stewart Lecker, Isaac Stillman *Boston University, Boston, MA:* Sushrut Waikar, *Brigham & Women’s Hospital, Boston, MA:* Gearoid McMahon, Astrid Weins, Samuel Short *Broad Institute, Cambridge, MA:* Nir Hacohen, Paul Hoover *Case Western Reserve, Cleveland, OH:* Mark Aulisio *Cleveland Clinic, Cleveland, OH:* Leslie Cooperman, Leal Herlitz, John O’toole, Emilio Poggio, John Sedor, Stacey Jolly *Columbia University, New York, NY:* Paul Appelbaum, Olivia Balderes, Jonathan Barasch, Andrew Bomback, Pietro A. Canetta, Vivette D. D’Agati, Krzysztof Kiryluk, Satoru Kudose, Karla Mehl, Jai Radhakrishnan, Chenhua Weng *Duke University, Durham, NC:* Laura Barisoni *European Molecular Biology Laboratory, Heidelberg, Germany:* Theodore Alexandrov *Indiana University, Indianapolis, IN*: Tarek Ashkar, Daria Barwinska, Pierre Dagher, Kenneth Dunn, Michael Eadon, Michael Ferkowicz, Katherine Kelly, Timothy Sutton, Seth Winfree *John Hopkins University, Baltimore, MD:* Steven Menez, Chirag Parikh, Avi Rosenberg, Pam Villalobos, Rubab Malik, Derek Fine, Mohammed Atta, Jose Manuel Monroy Trujillo, *Joslin Diabetes Center, Boston, MA*: Alison Slack, Sylvia Rosas, Mark Williams *Mount Sinai, New York, NY:* Evren Azeloglu, Cijang (John) He, Ravi Iyengar, Jens Hansen *Ohio State University, Columbus, OH:* Samir Parikh, Brad Rovin *Pacific Northwest National Laboratories, Richland, WA:* Chris Anderton, Ljiljana Pasa-Tolic, Dusan Velickovic, Jessica Lukowski *Parkland Center for Clinical Innovation, Dallas, TX:* George (Holt) Oliver *Patient Partners*: Joseph Ardayfio, Jack Bebiak, Keith Brown, Taneisha Campbell, Catherine Campbell, Lynda Hayashi, Nichole Jefferson, Robert Koewler, Glenda Roberts, John Saul, Anna Shpigel, Edith Christine Stutzke, Lorenda Wright, Leslie Miegs, Roy Pinkeney *Princeton University, Princeton, NJ:* Rachel Sealfon, Olga Troyanskaya *Providence Medical Research Center, Providence Health Care, Spokane, WA:* Katherine Tuttle *Stanford University, Palo Alto, CA:* Dejan Dobi, Yury Goltsev *University of California San Diego, La Jolla, CA:* Blue Lake, Kun Zhang *University of California San Francisco, San Francisco, CA:* Maria Joanes, Zoltan Laszik, Andrew Schroeder, Minnie Sarwal, Tara Sigdel *University of Michigan, Ann Arbor, MI:* Ulysses Balis, Victoria Blanc, Oliver He, Jeffrey Hodgin, Matthias Kretzler, Laura Mariani, Rajasree Menon, Edgar Otto, Jennifer Schaub, Becky Steck, Chrysta Lienczewski, Sean Eddy *University of Pittsburgh, Pittsburgh, PA:* Michele Elder, Daniel Hall, John Kellum, Mary Kruth, Raghav Murugan, Paul Palevsky, Parmjeet Randhawa, Matthew Rosengart, Sunny Sims-Lucas, Mary Stefanick, Stacy Stull, Mitchell Tublin *University of Washington, Seattle, WA:* Charles Alpers, Ian de Boer, Ashveena Dighe, Jonathan Himmelfarb, Robyn Mcclelland, Sean Mooney, Stuart Shankland, Kayleen Williams, Kristina Blank, Jonas Carson, Frederick Dowd, Zach Drager, Christopher Park *UT Health San Antonio, Center for Renal Precision Medicine, San Antonio, TX:* Kumar Sharma, Guanshi Zhang, Shweta Bansal, Manjeri Venkatachalam, *UT Southwestern Medical Center, Dallas, TX:* Asra Kermani, Simon Lee, Christopher Lu, Tyler Miller, Orson Moe, Harold Park, Kamalanathan Sambandam, Francisco Sanchez, Jose Torrealba, Toto Robert, Miguel Vazquez, Nancy Wang *Washington University in St. Louis, St. Louis, MO:* Joe Gaut, Sanjay Jain, Anitha Vijayan *Yale University, New Haven, CT:* Randy Luciano, Dennis Moledina, Ugwuowo Ugochukwu, Francis Perry Wilson, Sandy Alfano

## Supplementary Materials

### Supplementary Figures

**Supplementary Figure 1**: UMAP plot showing the distribution of LD and DKD cells in the 21 clusters

**Supplementary Figure 2**: Functional summary of DKD-induced ACE2+ expression signature.

**Supplementary Figure 3**: Heatmap of cluster markers used to identify cell types in COV

**Supplementary Figure 4**: ACE2 immunostaining in human kidney tissue from the Human Cell Atlas.

### Supplementary Tables

**Supplementary Table 1**: Clinical characteristics of COV cohort

**Supplementary Table 2**: Differential expression gene list for the LD ACE2+ signature.

**Supplementary Table 3**: Differential expression gene list for the DKD ACE2+ signature (Figure 4A)

**Supplementary Table 4**: Differential expression gene list for the DKD-induced ACE2+ signature genes (Supplementary Figure 2A)

**Supplementary Table 5**: The Gene Ontology enrichments for DKD ACE2+ signature network modules (Figure 4A).

**Supplementary Table 6**: The Gene Ontology enrichments for the DKD-induced ACE2+ signature network modules (Supplementary Figure 2A).

**Supplementary Table 7**: SARS-CoV-2 infection gene sets^20, 36, 37, 39^ with overlaps indicated for both the DKD ACE2+ signature genes and the COV ACE2+ signature genes (Supplementary Figure 2B)

**Supplementary Table 8**: A table of p-values indicating the significance of the genes shared between the Bojkova proteome^37^ gene list and each module of the DKD ACE2+ signature (Figure 4B).

**Supplementary Table 9**: A table of p-values indicating the significance of the genes shared between the Bojkova proteome^37^ gene list and each module of the DKD-induced ACE2+ signature (Supplementary Figure 2).

**Supplementary Table 10**: Cluster markers for single cell analysis of the COV samples (Figure 6).

**Supplementary Table 11**: Differential expression gene list for the COV ACE2+ signature (Figure 7A).

**Supplementary Table 12**: The Gene Ontology enrichments for network modules of COV ACE2+ signature (Figure 7A)

**Supplementary Table 13**: COV and DKD ACE2+ signature overlap gene set (Figure 7B)

**Supplementary Table 14**: The Gene Ontology enrichments for network modules of clustered genes shared by COV and DKD ACE2+ signatures (Figure 7B).

**Supplementary Table 15**: A table of p-values indicating the significance of the genes shared between the Bojkova proteome^37^ gene list and each module of the COV ACE2+ signature (Figure 7C).

**Supplementary Table 16**: The Gene Ontology enrichments for network modules for the network clustering of genes shared between the Bojkova proteome^37^ gene list and the COV ACE2+ signature (Figure 7C).

**Supplementary Table 17**: The Gene Ontology enrichments for network modules for the network clustering of genes shared between the Bojkova proteome^37^ gene list and the DKD ACE2+ signature (Fig 4B)

